# An updated open-source, large dataset of T1-weighted stroke brain MRIs and lesion segmentation masks

**DOI:** 10.64898/2026.11.25.26369004

**Authors:** Stuti Chakraborty, Mahir H. Khan, Ezequiel de la Rosa, Octavio Marin-Pardo, Bethany P. Tavenner, John Absher, Tim-Mathis Beutel, Felix Bitzer, Leonardo Bonilha, Michael R. Borich, Lara A. Boyd, Amy G. Brodtmann, Cathrin M. Buetefisch, Amelia Cain, Mariano Cabezas, Jessica M. Cassidy, Ashley Catchpole, Edward S. Chen, Steven C. Cramer, Julian Deseoe, Martin Domin, Adrienne N. Dula, Margarita Alethea Eidsness, Jennifer K. Ferris, Wuwei Feng, Emily E. Fokas, Niko H. Fullmer, Fatemeh Geranmayeh, Adrià Bermudo-Gallaguet, Makayla Gibson, Chris M. Gregory, Jeanette R. Gumarang, Leticia Hayes, Fabian von Hoch, Christina K. Holl, Brenton Hordacre, Abhishek Jaywant, Neda Jahanshad, Steven A. Kautz, Mohamed Salah Khlif, Jan S. Kirschke, Amisha Kumar, Kristan A. Leech, Sarah Lee, Jingchun Liu, Martin Lotze, Bradley J. MacIntosh, Maria Mataró, Bjoern Menze, Feroze B. Mohamed, Kyle Nishimura, Ava Pimentel, Fabrizio Piras, Nandita Raman, Shie Rinat, Kate P. Revill, Mauricio Reyes, Christopher Rorden, Emily R. Rosario, Heidi M. Schambra, Lena Schmitzer, Nicolas Schweighofer, Na Jin Seo, Grace C. Song, Surjo R. Soekadar, Ruisheng Su, Myriam Taga, Gregory T. Thielman, Sophia I. Thomopoulos, Paul M. Thompson, Ishaandeep Upadhyay, Daniela Vecchio, Lars T. Westlye, Benedikt Wiestler, Carolee J. Winstein, George F. Wittenberg, Kristin A. Wong, Chunshui Yu, Cansu Yalcin, Zhong Sheng Zheng, Sook-Lei Liew

## Abstract

Precise delineation of stroke lesions on MRI scans provides essential quantitative information used routinely in stroke rehabilitation research. Manual annotation continues to be the gold standard for stroke lesion delineation but is subjective and labor-intensive. Our previous release of ATLAS v2.1 (N=1271) provided a large, open-source dataset of T1w stroke brain MRIs and manually segmented lesion masks, which facilitated the development of numerous lesion segmentation algorithms. However, fully automated algorithm performance is not yet adequate to replace manual protocols; existing lesion segmentation algorithms still lack accuracy and generalizability. As model architectures grow increasingly sophisticated, they demand large-scale, diverse training data. Here, we present ATLAS v3.0 (N=1815), an updated, larger dataset of T1w stroke brain MRIs with corresponding manually segmented lesion masks. ATLAS v3.0 comes with nearly 43% more data, consisting of a publicly accessible, heterogenous training (n=1453), and a hidden generalizability dataset with subjects that remain unreleased (n=362). We anticipate this updated ATLAS v3.0 release will support the growing data demands of advanced lesion segmentation algorithms.

## Background & Summary

Stroke is a leading cause of death and disability worldwide.^1^ Brain imaging using non-invasive modalities such as Magnetic Resonance Imaging (MRI) and Computed Tomography (CT) have become invaluable tools to support timely diagnosis by clinicians, define optimal treatment strategies, and evaluate prognostic outcomes based on stroke etiology.^2^ Delineation of stroke-affected brain regions on imaging modalities (such as MRI scans) through lesion segmentation can provide important downstream spatial and quantitative information used in stroke rehabilitation research (e.g., tissue type classification and lesion load calculation, often complementing behavioral measures).^3^ Stroke lesion delineation can be performed either manually by trained annotators,^4^ or automatically by using trained algorithms. Manual annotation of stroke lesions remains the gold standard as it involves a trained annotator performing a thorough visual review to capture lesions of various sizes and patterns. However, it is a subjective and labor-intensive process, requiring annotators to delineate lesions on individual brain MRI slices one at a time, or using semi-automated (e.g., filling) techniques with manual correction.^4^ Automated methods are less time-consuming and more scalable, but face challenges in accurately delineating stroke lesions in particular.^3,5,6,7,8^ Some unique, stroke-specific challenges include the heterogeneity in the appearance of lesions depending on the post-stroke phase (e.g., acute, subacute or chronic) and type of imaging modality used (e.g. imaging, CT;T1-,T2-, or diffusion-weighted brain MRI), as well as the presence of other confounding pathologies and brain abnormalities that may be confused for stroke lesions.^3, 9^ Additionally, stroke lesions often appear irregular and have poorly defined boundaries; distinct lesions may sometimes appear merged together or blurred on imaging.^3^

To address these challenges, and to contribute to the development of accurate lesion segmentation algorithms, the ENIGMA Stroke Recovery Working Group has released some of the largest open-source datasets of T1-weighted (T1w) stroke brain MRIs with manually segmented lesion masks, including ATLAS v1.2 in 2018^10^ and ATLAS v2.1 in 2022.^9^ The ATLAS datasets have served as a key public benchmark, offering diverse and comprehensively annotated stroke lesion masks.^3,11,12^ Both these datasets have been used to develop numerous automated lesion segmentation algorithms for nearly a decade using various machine learning and more recently, deep learning architectures.^13–21^ ATLAS v2.1 has been cited more than 270 times and downloaded upwards of 3600 times by trainees, faculty, and the research community globally (as of September 2026).^22^

Notably, since the release of ATLAS v2.1, artificial intelligence (AI), and especially deep learning (DL) approaches, have undergone rapid transformations; this has resulted in marked improvements in medical image segmentation, as well as stroke lesion segmentation specifically.^23^ Several research-based and translational clinical applications of the ATLAS v2.1 dataset leveraging AI and DL advancements are reported in the literature.^24–26^ ATLAS v2.1 has continued to serve research purposes as a large-scale annotated dataset for algorithms, especially U-Net model architectures, used in automated stroke lesion segmentation. Importantly, ATLAS v2.1 has been used to develop “SynthSR”,^24^ an AI-driven tool that creates synthetic high-resolution (e.g., 1 mm isotropic) images from brain scans of any contrast and resolution, with the goal of overcoming sample size and power limitations in neuroimaging research while achieving spatial resolutions beyond those attained by conventional clinical scans.^24^ Using ATLAS v2.1 data has enabled SynthSR to account for stroke lesions that would have otherwise prevented downstream analysis (e.g., due to spatial normalization), when upsampling clinical brain MRIs. Other research-based uses of ATLAS v2.1 have included the pretraining of newer self-supervised learning, or foundation, models^25^ to capture innate, generalizable brain MRI representations,^26^ and to serve as an evaluation target to test the robustness of anomaly detection models trained on healthy brain scans.^19,27,28,29^ Beyond research-based applications, the ATLAS v2.1 dataset has also been used to perform clinically relevant downstream prediction tasks, such as in time-to-stroke predictions, which could enable clinicians to optimize time-sensitive interventions for stroke patients that may otherwise be excluded due to unknown stroke onset information.^19^ Despite this progress, the primary goal of the ATLAS v2.1 dataset release, to facilitate the development of robust and generalizable stroke lesion segmentation algorithms that match the performance of gold standard manual tracing, remains to be achieved at the time of this release.

Below, we summarize our review of some of the current best performing automated stroke lesion segmentation algorithms that were trained on ATLAS v2.1 data (Table 1), with the caveat that, given the growth in this area, we are unable to review all methods and cannot guarantee the accuracy of all methods. This table is primarily meant to provide a sample of current high-performing methods reported using ATLAS v2.1 to train different models, which move the field beyond the original scope of ATLAS v2.1. The dataset usage has addressed increasingly targeted challenges in stroke lesion delineation, including (i) achieving gains in segmentation performance on small lesions using multi-size and boundary aware labels^30^ or combining spatial and contextual features on top of an encoder-decoder architecture,^31^ (ii) delineating irregular boundaries of stroke lesions using an enhanced fuzzy logic to address ambiguous lesion appearances^32^, (iii) reducing image intensity disparities using a foreground harmonization framework to improve data augmentation,^33^ and (iv) evaluating the impact of the stroke lesion on focal descending motor pathways by calculating lesion volume and lesion load, in addition to automated segmentation.^34^ Our review, however, also reveals that existing methods have yet to meet our proposed Dice Similarity Coefficient [a measure of the overlap between the ground truth lesion and predicted masks (DSC)] cut-off of 0.85, to become an accurate, reliable replacement for manual segmentation.^4,9^ The *average* DSC across the best performing methods we identified is 0.691 (Table 1), which, although does not reach our proposed cut-off, could be adequate to use with a human-in-the-loop (HITL) system. In a HITL system, the algorithm segments first, and a human reviews and manually corrects lesions as needed. In our recent work, we have shown that this type of HITL system leads to a 47.4% reduction in time needed to annotate the lesion, compared to a fully manual process, with comparable accuracy^35^ (see *Identifying and tracing lesions* below).

**Table 1.**
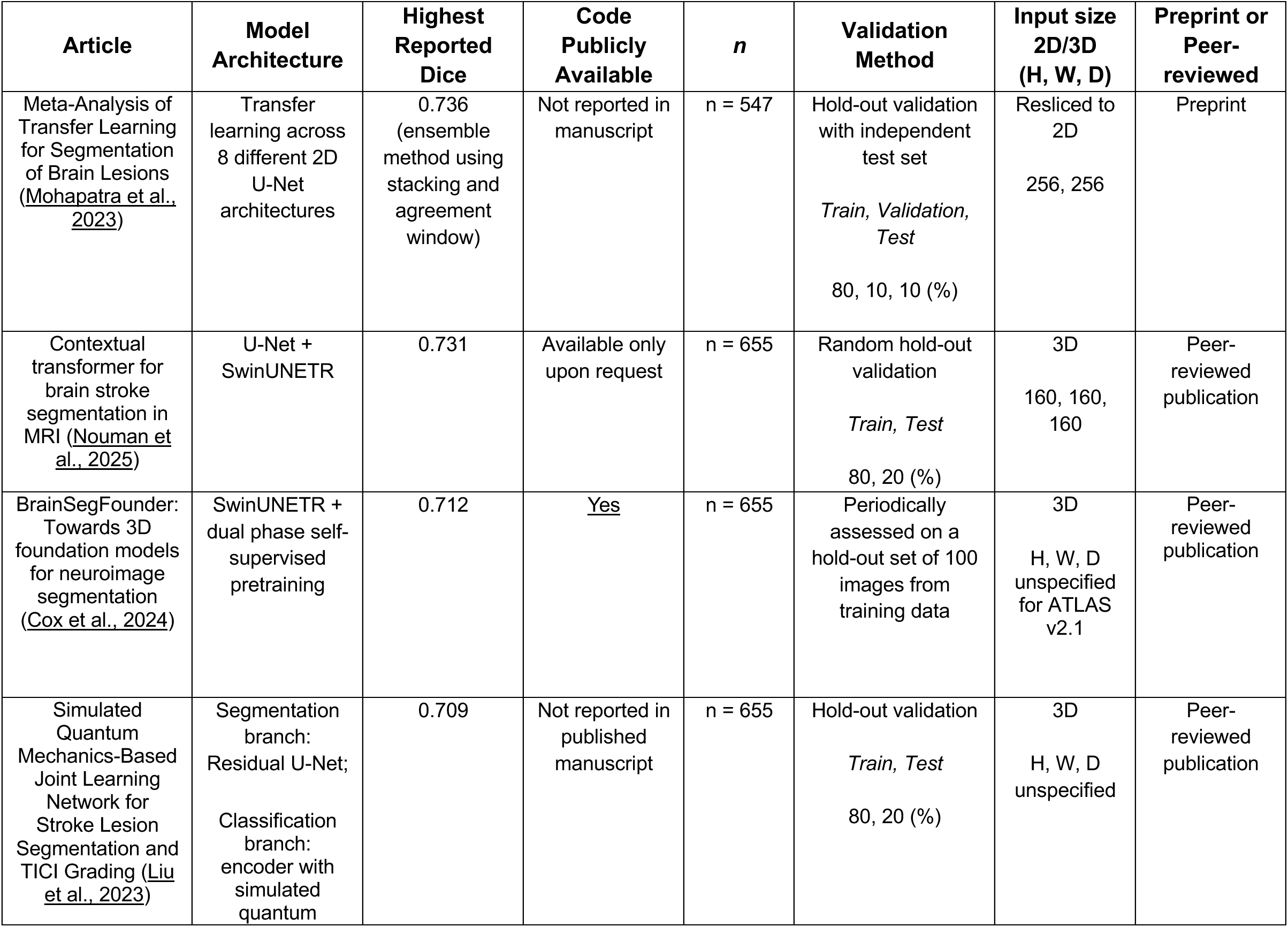
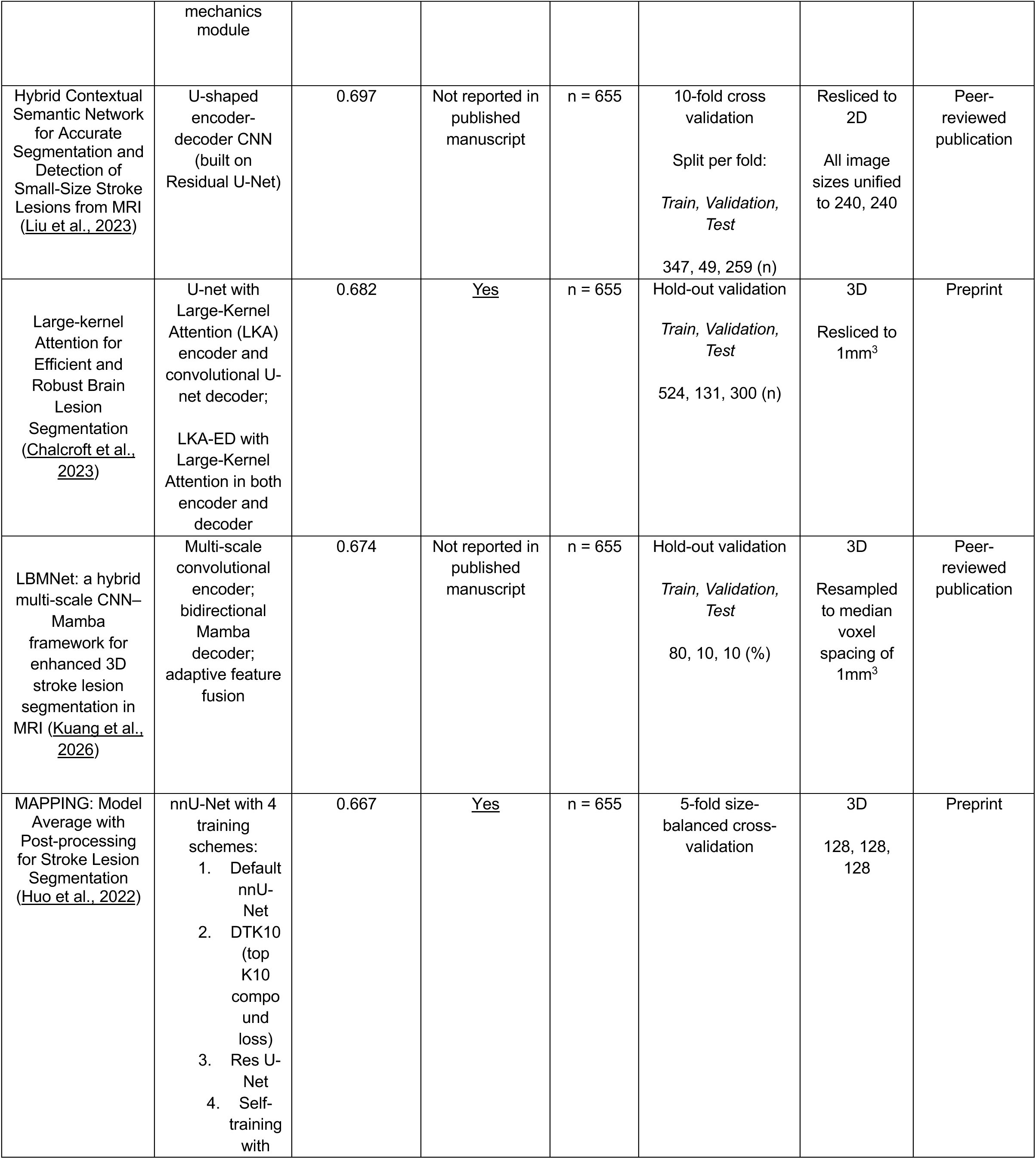
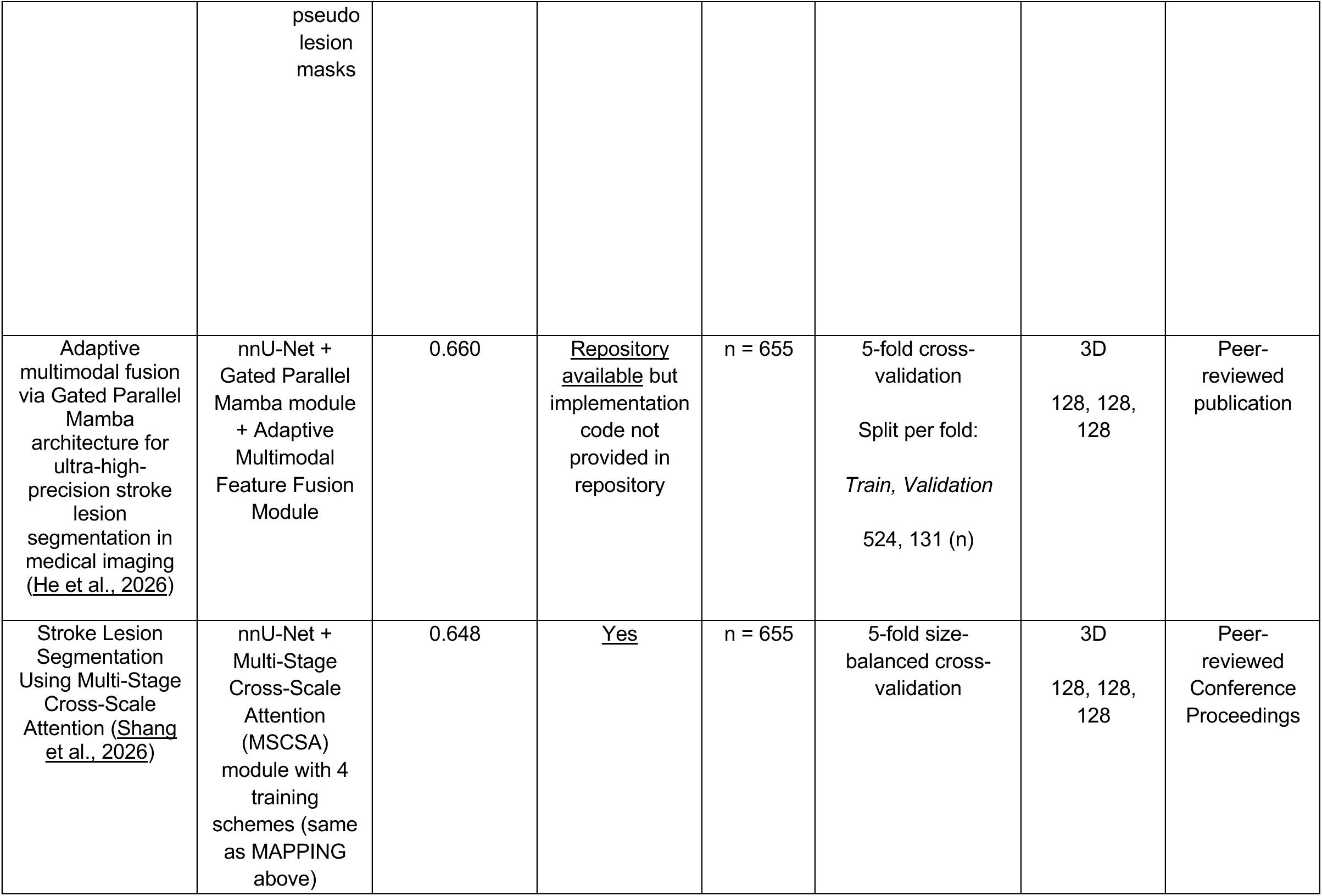
A Sample of High-Performing Published Methods for Automated Lesion Segmentation Using ATLAS v2.1. A summary of peer-reviewed publications/conference proceedings and preprints outlining automated lesion segmentation methods that were trained using ATLAS v2.1 data with brief notes on model architecture, highest reported DSC, code availability, subjects used (n), validation method and input image parameters. The automated lesion segmented method with the highest reported DSC coefficient is reported at the top, with the remaining organized in descending order of reported DSC.

| Article | Model Architecture | Highest Reported Dice | Code Publicly Available | *n* | Validation Method | Input size 2D/3D (H, W, D) | Preprint or Peer-reviewed |
|---|---|---|---|---|---|---|---|
| Meta-Analysis of Transfer Learning for Segmentation of Brain Lesions (Mohapatra et al., 2023) | Transfer learning across 8 different 2D U-Net architectures | 0.736 (ensemble method using stacking and agreement window) | Not reported in manuscript | n = 547 | Hold-out validation with independent test set<br>*Train, Validation, Test*<br>80, 10, 10 (%) | Resliced to 2D<br>256, 256 | Preprint |
| Contextual transformer for brain stroke segmentation in MRI (Nouman et al., 2025) | U-Net + SwinUNETR | 0.731 | Available only upon request | n = 655 | Random hold-out validation<br>*Train, Test*<br>80, 20 (%) | 3D<br>160, 160, 160 | Peer-reviewed publication |
| BrainSegFounder: Towards 3D foundation models for neuroimage segmentation (Cox et al., 2024) | SwinUNETR + dual phase self-supervised pretraining | 0.712 | Yes | n = 655 | Periodically assessed on a hold-out set of 100 images from training data | 3D<br>H, W, D unspecified for ATLAS v2.1 | Peer-reviewed publication |
| Simulated Quantum Mechanics-Based Joint Learning Network for Stroke Lesion Segmentation and TICI Grading (Liu et al., 2023) | Segmentation branch: Residual U-Net;<br>Classification branch: encoder with simulated quantum mechanics module | 0.709 | Not reported in published manuscript | n = 655 | Hold-out validation<br>*Train, Test*<br>80, 20 (%) | 3D<br>H, W, D unspecified | Peer-reviewed publication |
| Hybrid Contextual Semantic Network for Accurate Segmentation and Detection of Small-Size Stroke Lesions from MRI (Liu et al., 2023) | U-shaped encoder-decoder CNN (built on Residual U-Net) | 0.697 | Not reported in published manuscript | n = 655 | 10-fold cross validation<br>Split per fold:<br>*Train, Validation, Test*<br>347, 49, 259 (n) | Resliced to 2D<br>All image sizes unified to 240, 240 | Peer-reviewed publication |
| Large-kernel Attention for Efficient and Robust Brain Lesion Segmentation (Chalcroft et al., 2023) | U-net with Large-Kernel Attention (LKA) encoder and convolutional U-net decoder;<br>LKA-ED with Large-Kernel Attention in both encoder and decoder | 0.682 | Yes | n = 655 | Hold-out validation<br>*Train, Validation, Test*<br>524, 131, 300 (n) | 3D<br>Resliced to 1mm<sup>3</sup> | Preprint |
| LBMNet: a hybrid multi-scale CNN–Mamba framework for enhanced 3D stroke lesion segmentation in MRI (Kuang et al., 2026) | Multi-scale convolutional encoder; bidirectional Mamba decoder; adaptive feature fusion | 0.674 | Not reported in published manuscript | n = 655 | Hold-out validation<br>*Train, Validation, Test*<br>80, 10, 10 (%) | 3D<br>Resampled to median voxel spacing of 1mm<sup>3</sup> | Peer-reviewed publication |
| MAPPING: Model Average with Post-processing for Stroke Lesion Segmentation (Huo et al., 2022) | nnU-Net with 4 training schemes:<br>1. Default nnU-Net<br>2. DTK10 (top K10 compound loss)<br>3. Res U-Net<br>4. Self-training with pseudo lesion masks | 0.667 | Yes | n = 655 | 5-fold size-balanced cross-validation | 3D<br>128, 128, 128 | Preprint |
| Adaptive multimodal fusion via Gated Parallel Mamba architecture for ultra-high-precision stroke lesion segmentation in medical imaging (He et al., 2026) | nnU-Net + Gated Parallel Mamba module + Adaptive Multimodal Feature Fusion Module | 0.660 | Repository available but implementation code not provided in repository | n = 655 | 5-fold cross-validation<br>Split per fold:<br>*Train, Validation*<br>524, 131 (n) | 3D<br>128, 128, 128 | Peer-reviewed publication |
| Stroke Lesion Segmentation Using Multi-Stage Cross-Scale Attention (Shang et al., 2026) | nnU-Net + Multi-Stage Cross-Scale Attention (MSCSA) module with 4 training schemes (same as MAPPING above) | 0.648 | Yes | n = 655 | 5-fold size-balanced cross-validation | 3D<br>128, 128, 128 | Peer-reviewed Conference Proceedings |

Continued progress in DL model architecture development and prevention of model overfitting is dependent on large amounts of annotated training data.^23^ Concurrently, deployment of successful DL models is also dependent on their ability to adapt to various sources of MR image variability such as differences in scanner types, acquisition parameters and protocols.^3,35,36^ Such variability remains an inevitable aspect of real-world clinical datasets that must be adequately considered to ensure the training of robust, generalizable models.^36^ More recent model architectures such as transformer-based or hybrid CNN-transformers are better-positioned to be generalizable as they are able to: (i) incorporate textural patterns (using CNN capabilities), (ii) capture relationships between voxels that are far apart (long range dependencies), and (iii) contextualize global information, thereby gathering a more rigorous representation of shape-based attributes of lesions (using transformers).^12^ However, these architectures tend to exhibit suboptimal performance in stroke lesion segmentation when compared to brain tumors, due to the immense variability in infarct size, shape, location and other presentation of stroke lesions.^12,38^ Moreover, effective implementation of such advanced models is again dependent on large, heterogenous, multi-institution training data.^39,37^ Expectedly, the top performing segmentation methods trained on ATLAS v2.1 (Table 1) adopted image augmentation techniques to introduce variation in original images or to simulate synthetic disruptions in structural anatomy.^34,40^ Various other data augmentation techniques including spatial transformations and intensity perturbations^41^ have also been proposed.^42^ While data augmentation techniques can improve model generalizability and accuracy by introducing additional variability into existing images, they cannot fully replicate the diversity of stroke lesion locations, sizes, and anatomical patterns captured by real patient data, underscoring the continued importance of expanding both the size and heterogeneity of training datasets like ATLAS to improve lesion segmentation performance.

To address the evident need for larger, more diverse training data for enhancing automated lesion segmentation algorithms, here, we present the ATLAS v3.0 dataset, containing 1,815 T1w stroke brain MRIs and accompanying manual lesion segmentations from 61 research cohorts across 10 countries. The release of ATLAS v3.0 comes with nearly 43% more data, replacing and expanding upon ATLAS v2.1. ATLAS v3.0 enhances valuable data properties of preceding ATLAS versions,^9,10^ including: (i) sizeable heterogeneity in lesion location across diverse brain regions, (ii) diversity in lesion size across research cohorts, representing a spectrum of multiple smaller, punctuate lesions to large lesions spanning various anatomical structures, and (iii) further variations in data acquisition protocols, sources and scanner parameters (*see Data Characteristics and Supplementary Material, Tables 1 – 3; Figure 1)*.

ATLAS v3.0 is intended to continue to support the technical growth in DL model architectures discussed above. Another key motivation for the release of ATLAS v3.0 is to support the translational applicability of segmentation methods to heterogeneous lesion contexts for enhanced research and clinical utility across the stroke recovery continuum. To that end, we retain our focus on providing T1w stroke brain MRIs, with data primarily from the subacute and chronic phases of stroke.^9,10^ T1w imaging was strategically chosen due to several important reasons. First, although diffusion weighted imaging (DWI) is the gold standard for visualizing the extent of acute stroke damage,^43^ T1w images have been more commonly used in the subacute and chronic phases for post-stroke rehabilitation research to inform the prediction of functional outcomes.^44,45,46^ Second, unlike DWI, T1w images preserve lesion visibility across varied timepoints and can be used to better characterize structural changes in the cortex with progressive chronicity.^47,45^ Third, T1w images can be used to quantify post-stroke secondary sequelae (e.g., global atrophy^48–50^ and white matter hyperintensities)^51^ without the need for additional multimodal correction, which could be beneficial for longitudinal analyses. Fourth, newer hybrid CNN-Transformer architectures may be better suited to utilize the structural and anatomical granularity of T1w images, thereby enhancing the ability of CNNs to capture focal edge details with Transformers’ ability to contextualize global intra- or extra-lesion relationships.^12^ Finally, some recent approaches can be used to train a modality-agnostic model on T1w images (e.g., by generating augmented images using T1w MRIs that simulate other modalities), and subsequently test the trained model on real images from other modalities (DWI, T2, FLAIR, etc.), thus broadening the possible application of this work.^52^

Consistent with previous releases, the ATLAS v3.0 public training dataset has been released in two formats. First, we provide a minimally preprocessed dataset with native space images as a default to ensure algorithm developers can (i) select their own method of image normalization and (ii) develop methods that use as much or as little preprocessing. Additionally, this ensures preservation of uncompressed, original voxel dimensions and avoids any anatomical disruptions or loss of resolution that may be caused from registering images into MNI space. We have attempted to preserve original image properties to represent real-world data acquisition properties more closely, and have performed only one standardized, minimal preprocessing step (i.e. brain extraction to retain anonymity of participant data). Second, we also provide a preprocessed dataset, with brain-extracted images; all subjects in this dataset have undergone additional intensity normalization and registration to MNI space.

Notably, ATLAS v3.0 includes additional brain MRIs of stroke subjects with visible chronic lesions from the open-source Stroke Outcome Optimization Project (SOOP).^53^ SOOP is a retrospective dataset of MRI scans acquired during acute stroke care that also reveal chronic lesions from previous strokes. Stroke lesions in the SOOP data were originally traced on diffusion weighted images, however, lesion masks were subsequently coregistered to and manually corrected in T1w space to preserve the spatial resolution benefits provided by T1w images, essential for accurate lesion boundary delineation.^54^ This also enabled complementary information from both modalities to be used to determine lesion presence and extent, while maintaining consistency with the rest of the ATLAS dataset. Furthermore, we no longer retain the training/validation split of data from 955 subjects (training dataset = 655, validation dataset = 300) from ATLAS v2.1. Instead, we provide all 955 subjects as training data, including both T1w MRIs with lesion masks for all of these subjects. This should allow for wider validation of algorithms previously trained on ATLAS v2.1 and enable algorithm developers to choose their own validation approach. In addition, we have increased our total training data by 498 subjects (1453 from 955). Finally, we continue to retain the hidden generalizability dataset from ATLAS v2.1 and have added 46 new subjects from an unseen, independent research cohort to this dataset. This was done to increase the number of subjects available to test the generalizability of algorithm performance on unseen data (e.g., in the ISLES 2026 Challenge; see *Data Characteristics* below). Together, we anticipate that these enhancements will lead to the development of more improved lesion segmentation methods and enable robust research using our large, diverse dataset of stroke brain MRIs.

## Methods

### Data Overview

The complete ATLAS v3.0 dataset (including training and hidden generalizability) was aggregated from 61 research cohorts, with the addition of 17 new research cohorts since the ATLAS v2.1 release. As previously outlined, data included in ATLAS releases are generally collected for specific research purposes based on predetermined eligibility criteria. Hence, data contained in ATLAS v3.0 may not be representative of the general stroke population. Data were primarily collected to study mechanisms driving brain-behavior relationships in relation to sensorimotor, cognitive, speech-language, or mood-related impairments.^9^ Some of these data may have also been collected to investigate various post-stroke interventions.^9,10^ While a few observational or intervention studies collected longitudinal data, we have only included the first timepoint in ATLAS v3.0, in line with previous ATLAS version releases (see also *Data Records*). Data include acute (i.e., within the first 24 hours to 7 days after stroke), subacute (≥7 to 90 days), late subacute (≥90 to 180 days) and chronic (≥180 days after stroke) phases of stroke.^55^ Like ATLAS v1.2 and v2.1, data were derived from studies approved by respective local ethics committees and were conducted in accordance with the 1964 Declaration of Helsinki. Prior informed consent was obtained from all subjects. The ethics committee at the receiving research cohort (the University of Southern California) approved the receipt and sharing of the de-identified data, which did not contain any personal identifiers.

We performed quality control of brain MRIs contained within each subject file. Any images with large motion, ringing artifacts, or other distortions which may have obstructed lesion identification, were subsequently removed. All attempts were made to include as much data as possible. All subjects are named and stored in accordance with the Brain Imaging Data Structure (BIDS) (http://bids.neuroimaging.io/);^56^ subject identifiers are denoted in the metadata (column: ‘SESSION_ID”). The ATLAS v3.0 training dataset contains all the subjects included in ATLAS v2.1; information regarding whether a subject was included in prior releases is included in the metadata (column: “ATLAS2_DATASET”). For each subject, we also provide metadata on MRI acquisition time in relation to stroke onset (denoted as “DAYS POST STROKE”) if this data was made available by the contributing research cohort. In accordance with ATLAS v2.1 metadata, if information on days post stroke was not recorded or provided by the contributing cohort, a general timeline was used, denoted in the column labeled “CHRONICITY”. For example, if MRI was acquired at ≥180 days after stroke for a given subject, the chronicity column denotes a value of ‘1’, indicating that the subject is in the chronic phase, whereas values of 0 indicate non-chronic subjects. Although we have tried to provide as much data as possible, information on days post stroke or chronicity was not available for 3 research cohorts. The subject-level metadata also include the research cohort for each individual subject (denoted as “SITE”).

At the cohort-level, we provide a cohort-specific breakdown of the number of subjects included in the training dataset, along with the institution and country where the data were acquired, available in the *Supplementary Material (Supplementary Table 1)*. We also provide median days post stroke/chronicity by cohort and cohort-specific image acquisition parameters (for cohorts where complete image acquisition data were available) in the *Supplementary Material (Table 2 and Figure 1)*.

It is important to note that in upholding data sharing and privacy policies at several contributing cohorts, we do not release subject demographic information (e.g., age, sex), the presence of comorbidities or functional stroke outcomes.^9^ Where allowed, we aim to make demographic and clinical outcome data publicly available for a subset of our data in future releases.

### Data Characteristics

Similar to the ATLAS v2.1 dataset, ATLAS v3.0 consists of T1w MRI data collected on 1.5-Tesla and 3-Tesla MR scanners. The full training dataset for ATLAS v3.0 (n=1453) is derived from 55 distinct research cohorts. High-resolution data (i.e. voxel dimensions = 1mm^3^ isotropic) was available for all subjects in the training dataset from 36 research cohorts. The SOOP data also had subjects with ultra-high-resolution images (i.e. image dimensions <1mm^3^). Two cohorts (49 and 50) had subjects with all 3 voxel dimensions above 1mm, whereas 17 research cohorts had subjects with at least 1 voxel dimension above 1mm. A detailed summary of subject distribution by image dimension characteristics for each research cohort is provided in the *Supplementary Material* (*Supplementary Table 2)*. The generalizability dataset with hidden test data (n=362) adds 46 subjects from a new research cohort, resulting in data from a total of 12 research cohorts. We hope that this data will continue to be used to validate the accuracy and performance of trained algorithms on unseen subjects.

Although this information was not made available with the public release, here, we report aggregated metadata on the number of lesions, the lesioned hemisphere (e.g., left vs. right or bilateral), anatomical location (e.g. cortical or subcortical vs. brainstem or cerebellum) across subjects included in the training dataset (n=1453). Additionally, we have provided a cohort-level breakdown of the median lesion volumes of subjects included in the training dataset, in MNI space (*Supplementary Material, Table 3*). We anticipate that this detailed information made available for each subject will facilitate data sorting for further subgroup analyses and algorithm training based on diverse lesion characteristics.

From the training dataset (n=1453), 1384 subjects had data available on days post stroke or chronicity. Although we have attempted to maximize information on days post stroke or chronicity, this information was not available for 69 subjects. Of the 1384 with data available, 87 (6%) subjects were in the acute, 261 (18%) subjects were in the subacute, 115 (7.9%) subjects were in the late subacute, and 921 (63.4%) subjects were in the chronic phase of stroke. For the complete training dataset (n=1453), the median [IQR (Q1 – Q3)] lesion volume (in MNI space) was 7346 [46,418(1542 – 47,960)] mm^3^.

We used our Pipeline for Analyzing Stroke Lesions (PALS)^57^ toolbox to calculate lesion voxel overlaps with FreeSurfer Regions of Interest (ROIs); these outputs were subsequently summarized to obtain a group-level overview of the lesioned hemisphere, affected anatomical region (e.g. cortical/subcortical/brainstem/cerebellar) and number of lesions in the affected anatomical regions, for subjects included in the training dataset (n=1453). Five subjects had no visible lesions. The lesioned hemisphere for all subjects in the training dataset with visible lesions (n=1448) is reported in Table 2. 536 subjects (37.01%) had one lesion, of which, a majority had lesions in the right hemisphere [n=255 (17.61%)]. 912 (62.98%) subjects had multiple lesions, of which a majority had bilateral hemispheres affected [n=487 (33.63%)]. The number of lesions by anatomical region is reported in Table 3.

**Table 2.** Lesioned hemisphere for subjects in the training dataset (n = 1453) Subjects with a single, and multiple lesions, classified into left, right and bilateral hemispheres are reported. For subjects with a single lesion classified as bilateral, lesions extended into voxels overlapping with regions of interest (ROIs) across both hemispheres. Five subjects had no visible lesion (0.03%). Percentages are reported based on the total subjects with visible lesions (n = 1448) in the training dataset.

| **Subjects with One Lesion** |  |  | **Subjects with Multiple Lesions** |  |  |
|---|---|---|---|---|---|
| Left | Right | Bilateral | Left | Right | Bilateral |
| 229<br>(15.81%) | 255<br>(17.61%) | 52<br>(3.59%) | 210<br>(14.50%) | 215<br>(14.84%) | 487<br>(33.63%) |

**Table 3.** Number of lesions by anatomical region for subjects in the training dataset outputs (n = 1453) The number of lesions identified within specific regions (e.g. cortical, subcortical, brainstem and cerebellar) separated by hemisphere, is shown for subjects included in the training dataset (n = 1453). Note that it is possible for the total number of lesions to exceed the total number of subjects, as several subjects have more than one lesion. A subject could have lesions with voxel overlaps across multiple ROIs thus adding to the count, e.g. a subject could have both a cortical and cerebellar lesion.

| **Lesion Location** |  |  |  |  |  |  |  |
|---|---|---|---|---|---|---|---|
| **Cortical** |  |  |  | **Subcortical** |  |  |  |
| Left | Right | Bilateral | Total | Left | Right | Bilateral | Total |
| 463 | 443 | 416 | 1322 | 392 | 451 | 235 | 1078 |
| **Brainstem** |  |  |  | **Cerebellar** |  |  |  |
| Left | Right | Bilateral | Total | Left | Right | Bilateral | Total |
| 131 | 119 | 152 | 402 | 90 | 87 | 45 | 221 |

Maintaining consistency with the ATLAS v2.1 data release, we continue to not provide any metadata for the hidden generalizability dataset reported in ATLAS v2.1 (n=316), or the subjects added in ATLAS v3.0 (n=46). The hidden generalizability dataset is designed to validate algorithms on both in-sample and out-of-sample subjects; this was done by including held-out subjects from cohorts in the training sample, as well as from completely independent, unseen cohorts. Only participants of lesion segmentation challenges may have the hidden generalizability dataset available for evaluation (see *Lesion Segmentation Challenges* below).

### Training for individuals performing lesion tracing

Lesion tracing was performed using ITK-SNAP (version 4.2.2) by trained research assistants (tracers) following standardized protocol^4^ developed by the same team that created ATLAS v1.2^10^ and v2.1.^9^ We included additional improvements in the lesion training process to ensure sustained quality in segmentation outputs. Tracers received further in-depth training through narrated tutorials on neuroanatomy, distinguishing between MR imaging modalities and identifying stroke pathology around challenging regions (e.g., the insula). These narrated tutorials are publicly accessible on YouTube under the playlist *“Neuroanatomy for Lesion Tracing Training”* (https://youtube.com/playlist?list=PLPqz6WxiWPVE&si=07x053k2XGQmTpb4).

Specific tracing guidance that focused on in-depth distinction of lesions from commonly mis-segmented structures such as perivascular spaces, lacunae, and white matter hyperintensities were introduced^58^ (see *Identifying and tracing lesions*). Our lesion tracing guidance document has been made publicly accessible via GitHub (https://github.com/npnl/ATLAS/tree/master/Lesion%20Tracing%20Training). Our standard operating procedures have been previously published,^4^ and our lesion training datasets are publicly available for access (https://github.com/npnl/ATLAS). The training module for medical image segmentation using ITK-SNAP^59^ is available freely (http://www.itksnap.org/docs/fullmanual.php) and was recommended reading as part of the lesion tracing process.

### Identifying and tracing lesions

We followed the same lesion identification and tracing steps as previously outlined for the ATLAS v2.1^9^ dataset for all subjects. For subjects in one cohort (i.e. SOOP), segmented lesion masks were made available by the contributors; trained ATLAS team members carefully reviewed and corrected lesion boundaries following our standardized manual lesion segmentation protocol.^4^ This was done to ensure lesion masks were made as uniform as possible across the entire ATLAS v3.0 dataset. Briefly, tracers opened each individual T1w MRI in ITK-SNAP and carefully inspected them slice-by-slice, beginning at the posterior aspect of the brain (i.e., the cerebellum). Tracers were encouraged to scan for lesions across all 3 image planes (axial, coronal, sagittal), but trace only on the axial or coronal views.^10^ Lesion tracing was performed using a stylus (e.g., on an Apple iPad) or a mouse. All identified lesions from each individual subject were saved together as a single segmentation mask, using a BIDS compliant naming scheme (further details in *Data Records*).

Guidance on distinguishing between lesions and imaging-based markers of cerebral small vessel disease (CSVD; e.g., lacunes and white matter hyperintensities) was provided to tracers, in accordance with the STRIVE (Standards for Reporting Vascular changes on Neuroimaging) criteria^58^ and further reviewed by a trained neuroradiologist with >10 years of experience (G.B.). A lacune was identified as bigger than perivascular spaces (3-15 mm), asymmetric, with irregular shapes (often triangular). Tracers were prompted to pay closer attention to subcortical structures such as the basal ganglia nuclei where lacunae are commonly found (**Figure 1**). Perivascular spaces were noted to be symmetric, smaller than 3 mm, and with regular tubular shapes following the course of blood vessels (**Figure 1**). Periventricular white matter hyperintensities were noted to be contiguous with the borders of lateral ventricles. On the other hand, tracers were guided to check throughout the course of the white matter to locate deep white matter hyperintensities.^58^ Tracers were instructed to only capture stroke lesions in their annotation masks; CSVD markers were excluded.

**Figure 1:**
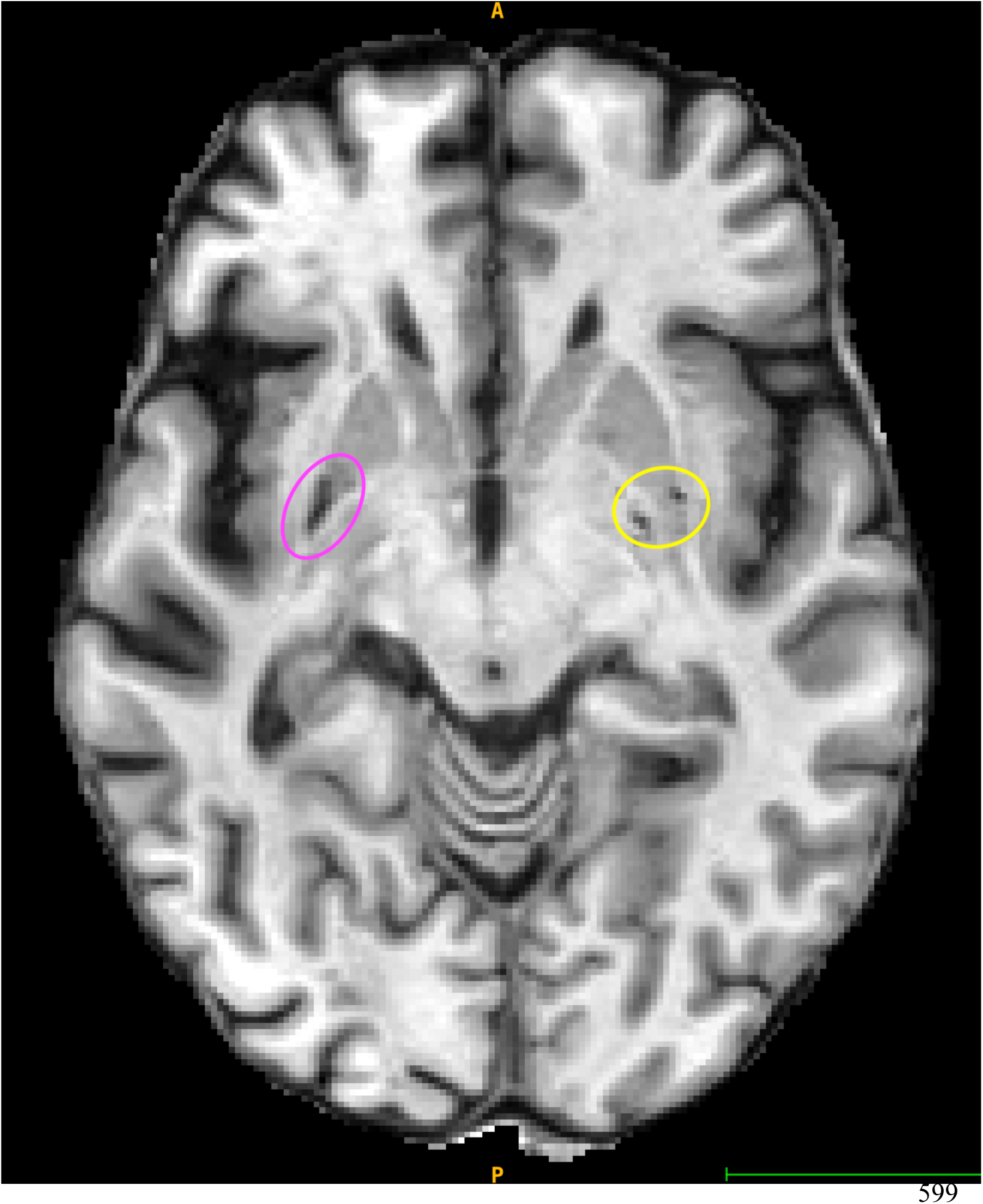
Example of lacunae and perivascular spaces included in lesion tracing training. An example of the ITK-SNAP software interface displaying an axial slice with a lacune, larger in size (circled in pink) and perivascular spaces, much smaller in size, with tubular appearance (circled in yellow).

For the last small subset of subjects in ATLAS v3.0 (n=66), we introduced a modification to our lesion segmentation workflow. The primary goal of this modification was to test whether automated lesion segmentation outputs generated by training a DL model with transformer-based implementation could be used as a starting point, thus reducing the time associated with manual tracing while still involving human review and correction for accuracy (i.e., by adopting a human-in-the loop (HITL) approach)^35^. Detailed information regarding model architecture, training parameters, and model performance have been described in detail by Khan et al.^35^ Briefly, we trained a publicly available nnU-Net model using ATLAS v2.1 data^35,60^ to automatically segment stroke lesions. This architecture was adopted for its ability to configure itself, including any training, pre- and post-processing steps involved, for new tasks and required limited manual intervention. Outputs generated by the automated segmentation method were subsequently corrected and quality checked for accuracy by two trained members with >3 years of manual lesion tracing experience (S.C. and M.H.K).

### Preprocessing normalization, registration and defacing

To reduce any possibilities of re-identification that may occur with traditional defacing approaches,^61^ we used SynthStrip,^62^ an open-source deep-learning based method available with FreeSurfer (https://surfer.nmr.mgh.harvard.edu/docs/synthstrip/), to perform a brain-only extraction, thus anonymizing each image. SynthStrip also enables more robust brain extraction for brains impacted by pathology such as stroke. As an update from prior ATLAS versions, for ATLAS v3.0 registration, we used SynthMorph,^63^ another open-source, deep-learning based method packaged with FreeSurfer to register images (https://martinos.org/malte/synthmorph/#tool). We implemented an affine-only registration (as opposed to a nonlinear warp) to the MNI152 template^64^; this was done to avoid the lesion biasing spatial normalization, thereby preserving its volume, shape, and boundaries, as well as adjacent anatomy. The segmented brain mask was then co-registered to template space using nearest neighbor interpolation. Advantages of SynthMorph include its ability to register brain images of any volume, size, resolution or contrast without the need for prior preprocessing. After preprocessing, we visually assessed whether each subject’s lesion mask registered properly to MNI space. Stroke lesion volumes (calculated as the total number of voxels within the lesion mask over anatomical regions of interest) were then obtained using our publicly available PALS toolbox (https://github.com/npnl/PALS). We note that the lesion volumes obtained from PALS were not released publicly with the training dataset (n=1453) and are only reported at a group-level across all subjects, in this paper. Details of the lesion tracing and preprocessing pipeline are summarized in **Figure 2**. A visual example of post-processing quality control is shown in **Figure 3**.

**Figure 2.**
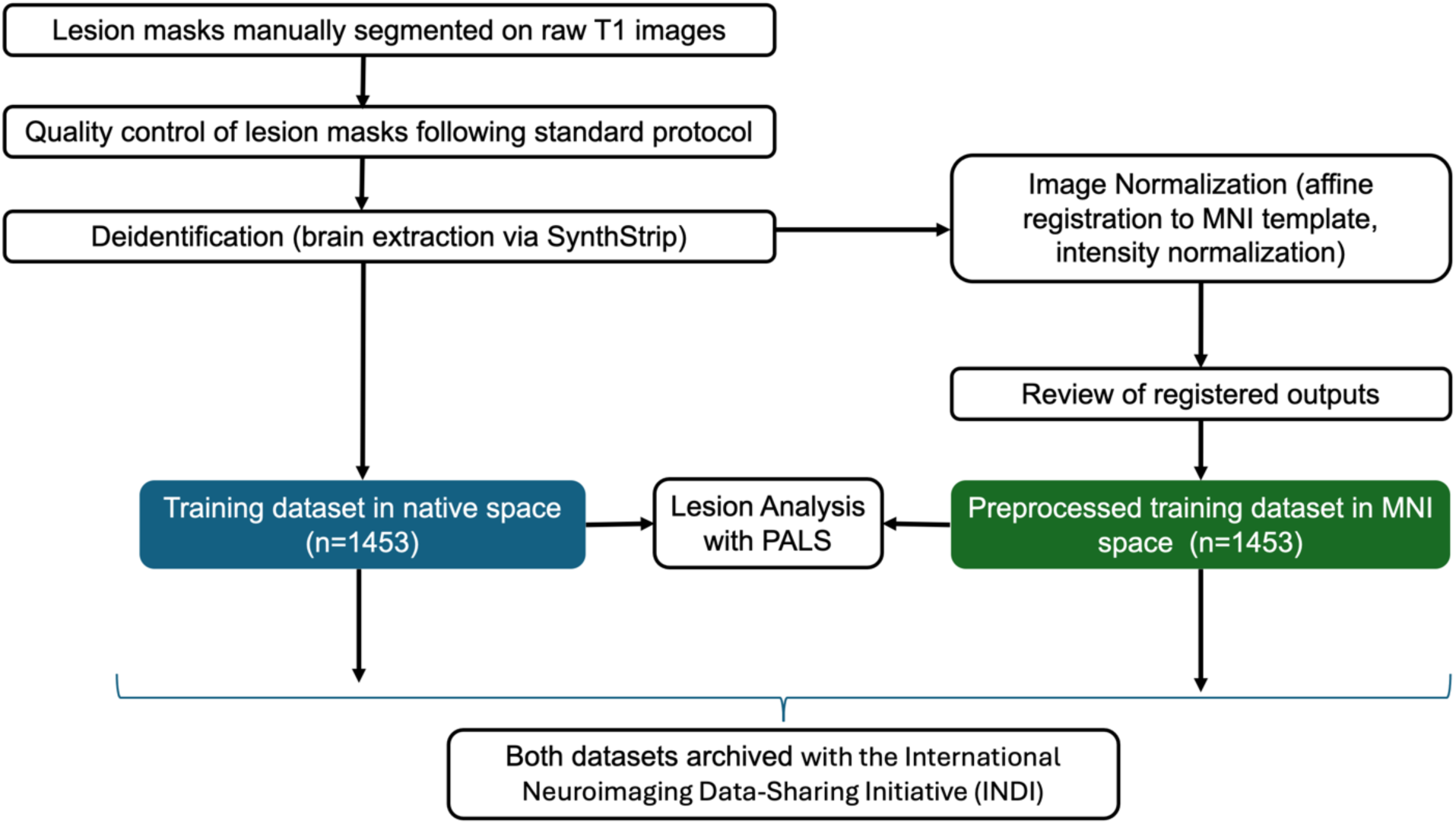
Lesion Tracing and Preprocessing Pipeline. A flow diagram representing the process of creating the ATLAS v3.0 publicly released training datasets (n=1453). The minimally processed (with brain extraction only) (left blue box) and the preprocessed (with additional affine registration to MNI template and intensity normalization) (right green box) datasets are archived with the International Neuroimaging Data-Sharing Initiative (INDI). Abbreviations: PALS = Pipeline for Analyzing Lesions After Stroke

**Figure 3.**
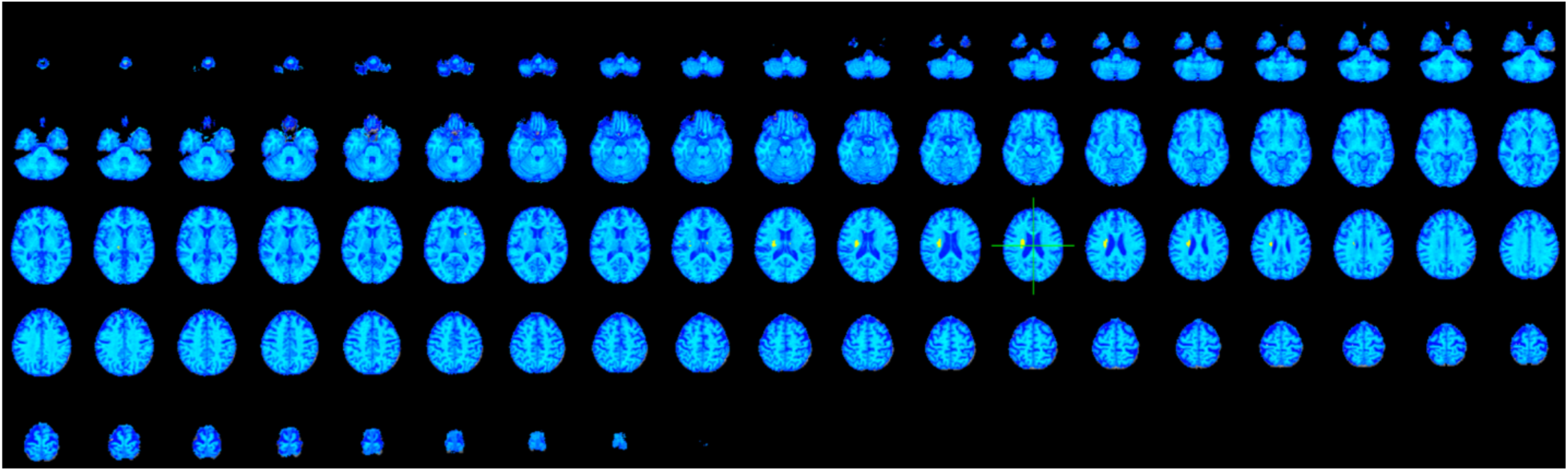
Post Preprocessing Visual Quality Control. Example of visual quality control after preprocessing to ensure proper registration of a stroke individual’s brain and segmented lesion mask (yellow), to the MNI template (blue) in the axial view, for a subject from the ATLAS v3.0 public training dataset.

### Probabilistic Spatial Heatmap of Lesion Location

We visualized the spatial distribution of lesions across all subjects publicly released as in the ATLAS v3.0 training dataset by summing the segmented lesion masks across all subjects in the MNI template space (**Figure 4**). Higher values indicate locations where lesions were present in a greater number of subjects.

**Figure 4.**
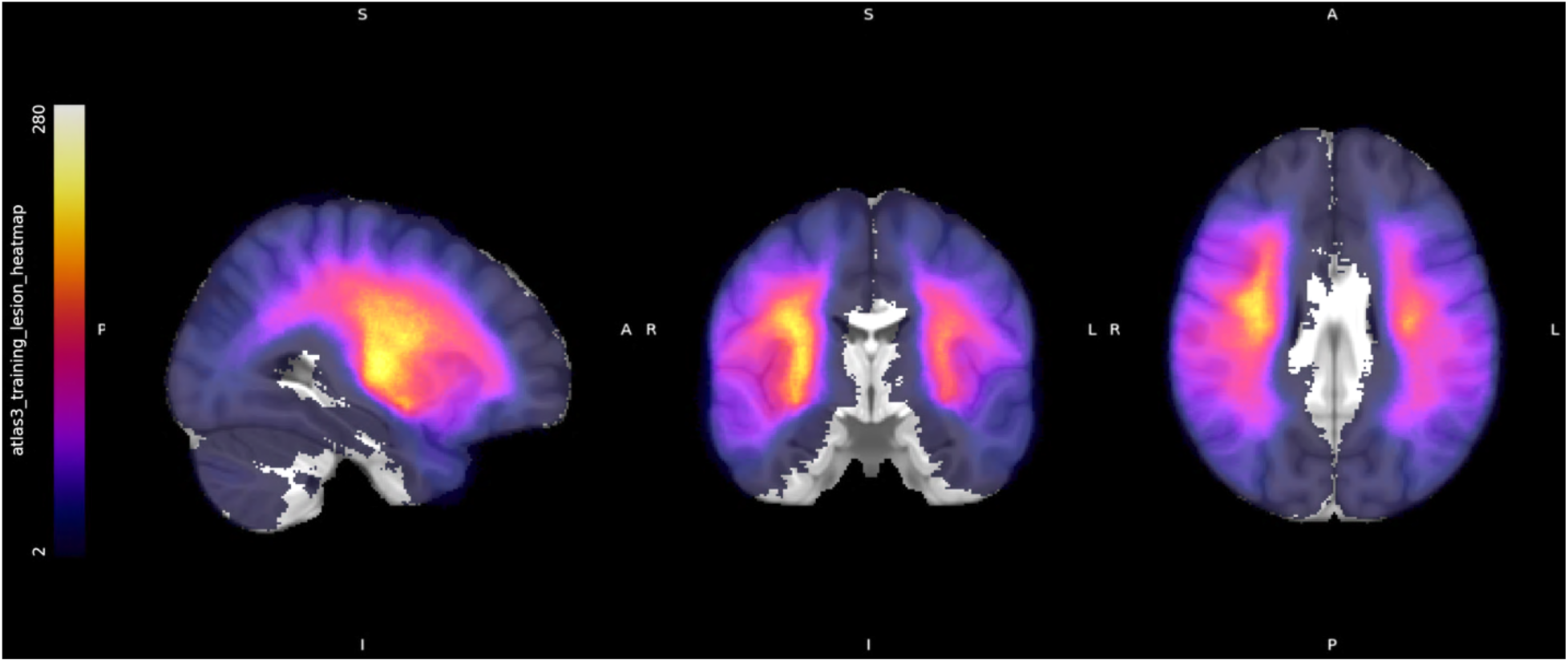
Probabilistic Lesion Overlap Map on the MNI_icbm152 Template. Lesion overlap visualization across all subjects in the ATLAS v3.0 public training dataset (n=1453) overlaid on the MNI template (brain-extracted). Warmer colors indicate greater number of subjects with lesions at that voxel.

#### Data Record

ATLAS v3.0 data are publicly available to download from the International Neuroimaging Data-sharing Initiative (INDI) https://fcon_1000.projects.nitrc.org/indi/retro/atlas_download.html. Within this link, ATLAS v3.0 training data are provided as both minimally processed (only brain extracted) T1w images in native space and preprocessed images. However, for the purposes of the most recent lesion segmentation challenge, only the use of native space T1w images was permitted (see *Lesion Segmentation Challenges* below). For ATLAS v2.1 since raw (non-brain-extracted) data were released, data were archived with the Archive of Data on Disability to Enable Policy and research (ADDEP),^65^ upholding strict data use agreements for data privacy reasons. Since all data in ATLAS v3.0 have undergone brain extraction, additional data archiving with ADDEP was not required.

All data are maintained following the Brain Imaging Data Standard (BIDS).^66^ We continue to provide individual subject folders within respective cohort folders. Each individual subject folder has a sub-folder named *“anat”,* which consists of a T1w MRI file, a corresponding lesion segmentation mask and an accompanying .csv metadata file containing relevant metadata (e.g. days post stroke, site, whether included in ATLAS v2.1 data). We follow the same *“sub-r***s***_ses-1”* naming convention as ATLAS v2.1, where *“r***”* corresponds to the research cohort number, *“s***”* corresponds to the subject number, and *“ses-1”* corresponds to a cross-sectional, single timepoint, for subjects from all research cohorts with the exception of the SOOP subjects. The SOOP subjects follow a slightly modified convention i.e. *“sub-soop****”*, where “soop****” corresponds to the subject number. We also note a few minor changes in the native space and preprocessed MNI space naming conventions. The minimally processed T1w MRI in native space are named *“sub-r***s***_ses-1_space-orig_desc-brain_T1w.nii.gz”*, and the corresponding lesion masks are named *“sub-r***s***_ses-1_space-orig_label-lesion_desc-T1lesion_mask.nii.gz”*. Consequently, the preprocessed T1w MRI in MNI space are named *“sub-r***s***_ses-1_space-MNI152NLin2009aSym_desc-brainnorm_T1w.nii.gz”* and the corresponding lesion masks in MNI space are named *“sub-r***s***_ses-1_space-MNI152NLin2009aSym_label-lesion_desc-T1lesion_mask.nii.gz”*.

#### Technical Validation

We used our standardized and previously published lesion segmentation protocol in the preparation of ATLAS v3.0; details of the technical validation parameters of our protocol have been previously published.^4^ Numerous automated lesion segmentation methods have been developed using the ATLAS v2.1 dataset, ^17,67,68,69,70,71^ several of which have been summarized above (Table 1, *Background and Summary*). We performed manual quality checks for all subjects included in the ATLAS v3.0 dataset; segmented lesion masks in the native space were quality controlled by trained team members following our standardized protocol. After preprocessing, lesion masks were examined for appropriate registration to the MNI template space.

#### Usage Notes

As noted for previous versions of ATLAS, data may be accessed under a standard Data Use Agreement, restricting data usage only for research (e.g., model training, testing) and statistical purposes. Data must not be used for the investigation of specific research subjects. Usage of the ATLAS v3.0 dataset, contributions of co-authors and their institutions should be appropriately acknowledged by citing this article and the specific data repository used for accessing this dataset. While we enhanced our standardized and previously published manual lesion segmentation protocol^4^ for this release, and continued robust quality control, we acknowledge that the manual segmentation process can be subjective. We will continue to review errors or issues in lesion segmentation, which can be submitted on the ATLAS GitHub page under ‘*issues*’ (https://github.com/npnl/ATLAS/). Modifications to the dataset or updates with new data will be released under new versions of ATLAS, as done previously. Any changes will be posted on the ATLAS GitHub.

Shortly before the release of ATLAS v3.0, we released MAESTRO, an open-source, generalizable model for lesion segmentation, briefly described above.^35^ MAESTRO exhibited robustness across diverse image inputs and significantly reduced annotation time when a human-in-the-loop workflow was adapted to correct automated lesion segmentations outputs from MAESTRO. MAESTRO is publicly accessible on our GitHub (see *Code Availability*).

### Lesion Segmentation Challenges

The provision of a hidden generalizability dataset to ensure fair performance evaluation of automated algorithms submitted to lesion segmentation challenges remains a key purpose of ATLAS v3.0, as done previously with ATLAS v1.2 and v2.1. Consistent with previous ATLAS dataset releases^9,10^, the Medical Image Computing and Computer Assisted Intervention (MICCAI) Ischemic Stroke Lesion Segmentation Challenge (ISLES) 2026 edition (https://isles-26.grand-challenge.org/) for ATLAS v3.0 was launched. With this iteration of the challenge, we hoped to address the out-of-domain generalizability gaps that emerged from ISLES 2022.

The ATLAS v3.0 training data for ISLES ‘26 was publicly released on April 24, 2026; the challenge concluded on August 19, 2026. The ISLES ‘26 challenge called for participants to develop automated stroke lesion segmentation algorithms using ATLAS v3.0; participants were ranked based on both technical proficiency and clinical impact. Performance evaluation metrics included the (i) DSC, which measures the overlap between the predicted and ground truth segmentation masks and normalizes the overlap based on the sizes of the combined regions, (ii) Absolute Volume Difference (a measure of the difference between the true and predicted lesion volumes), (iii) Absolute Lesion Count Difference (a measure of the difference between the number of ground truth and predicted lesions), (iv) Lesion-wise F1 score (which measures whether a singular voxel of a ground truth lesion is detected) and, (v) Area Under the Precision-recall Curve (which is computed as the weighted mean of precision values interpolated across operating points, given weightage based on the increase in recall).^72,73^ Further details on sample metrics can be found on GitHub (https://github.com/ezequieldlrosa/isles26/tree/main/utils). For this challenge, participants were encouraged to focus on algorithm development using only T1w images provided in the native coordinate system. The use of standardized or normalized versions of the data was not permitted.

Detailed information regarding the ISLES ‘26 challenge has been described by de la Rosa et al.^73^ Consistent with previous versions, this challenge was conducted in three phases.^74^ In the training phase, along with the first batch of training data, a Python evaluation script was released, which could be used to compute the above-mentioned performance evaluation metrics, enabling participants to test and iterate models accordingly. The sanity-check phase required participants to submit a sample docker to test their algorithm’s functionality on organizers’ servers. The test phase required participants to submit a docker containing their final algorithm submissions which were evaluated to declare rankings.^74^ We received 261 submissions for the Preliminary Docker Evaluation (sanity check) phase, and 65 submissions for the final test phase. Submissions for ISLES ‘26 were received from around the world, with the highest recorded submissions from China (60), India (53), United States (47), and Canada (14). The ISLES ‘26 official rankings and final performance metrics will be announced later in 2026.

The widespread use of the ATLAS datasets highlights their impact on the stroke research and medical image segmentation communities. To that end, we aim to continue to provide as much data as possible for training and improving lesion segmentation algorithms. As previously outlined, the ATLAS datasets are compiled by an international consortium of stroke researchers (ENIGMA Stroke Recovery Working Group)^75^, which routinely receives stroke brain MRI data from all over the world. Future ATLAS releases may incorporate clinically relevant neuroimaging markers of CSVD such as perivascular spaces and white matter hyperintensities, making them available along with lesion masks. This may aid in the development of algorithms sensitive to delineating pathology involving CSVD markers and reducing mis-segmentations.^76^ Further, successive lesion segmentation challenges may focus on developing automated segmentation methods that expansively capture relevant CSVD markers in addition to stroke lesions.

To conclude, here, we release ATLAS v3.0, which includes a total of 1815 T1w stroke brain MRIs, with 1453 containing corresponding lesion masks as training data, and a hidden generalizability dataset (n=362) which is made available only for lesion segmentation challenges. The primary goal of the ATLAS v3.0 dataset release is to better represent the heterogeneity of lesion presentation across stroke brain MRIs. A nearly 43% increase in data from ATLAS v2.1, our dataset captures several sources of variation, closely representing real-world data (including lesion size, anatomical location, time of MRI acquisition etc.). Additionally, we anticipate that as model architectures like transformers^12^ gain popularity, larger and more diverse datasets may be beneficial for effective model training and generalization, although human-in-the-loop approaches are likely to provide the optimal solutions.

## Data Availability

ATLAS v3.0 data are publicly available to download from the International Neuroimaging Data-sharing Initiative (INDI) https://fcon_1000.projects.nitrc.org/indi/retro/atlas_download.html. Within this link, ATLAS v3.0 training data are provided as both minimally processed (only brain extracted) T1w images in native space and preprocessed images. Stroke lesion segmentations were manually annotated using ITK Snap version 4.2.2. Lesion tracing training subjects from our lesion segmentation protocol^4^ can be found on GitHub (https://github.com/npnl/atlas). For the ISLES ‘26 lesion segmentation challenge, see https://isles-26.grand-challenge.org/. Instructions on downloading, decrypting and requesting password access to obtain the data can be found on the INDI website (https://fcon_1000.projects.nitrc.org/indi/retro/atlas_download.html).

## Code Availability

SynthMorph and SynthStrip are publicly available tools that can be found at https://martinos.org/malte/synthmorph/#tool and https://surfer.nmr.mgh.harvard.edu/docs/synthstrip/). The Pipeline for Analyzing Lesions after Stroke (PALS), our publicly available software to perform lesion load calculation and other image preprocessing, can be accessed from https://github.com/npnl/PALS.

MAESTRO, our automated lesion segmentation model trained on ATLAS v2.1 data, is available on our GitHub (https://github.com/npnl/MAESTRO).

## Author Contributions

This manuscript was read, edited, reviewed and approved by all listed authors. S.C. provided lesion segmentation training to tracers, oversaw the lesion segmentation process, traced lesions, wrote, and edited the manuscript. M.H.K. oversaw data intake, preparation, processing and edited the manuscript. M.H.K., E.D.L.R, M.C., J.D., B.M., J.S.K., M.R., C.R., L.S., B.W., R.S., and C.Y. oversaw the ISLES ‘26 challenge. O.M.P, B.P.T and M.H.K. traced and reviewed lesions and contributed to the preparation of the dataset. A.C., A.C., C.H., E.A.M., A.M.K, S.L., K.N., A.P., N.R., G.C.S., traced lesions. I.U. helped with data summarization. M.R.B., L.A.B., A.G.B., T.R.B., C.M.B., J.M.C., E.S.C, S.C.C., M.A.D., M.D., A.N.D., M.A.E., E.R.E., J.K.F., W.F., E.E.F., N.H.F., F.G., A.B., M.G., C.M.G., J.R.G., C.A.H., L.H., C.H., B.H., A.J., S.A.K., M.S.K., K.A.L., J.L., M.L., B.J.M., M.M., F.M., F.P., A.D.R., S.R., E.R.R., H.M.S., N.S., N.J.S., S.R.S., M.T., G.T.T., D.V., S.J.W., L.T.W., C.J.W., G.F.W., K.A.W., C.Y., Z.S.Z and S-L.L. were involved in data acquisition, deidentification and sharing of data released in the ATLAS v3.0 dataset. N.J., S.I.T., and P.M.T., provided support for conceptualization of the dataset. S-L.L. conceptualized the dataset, oversaw data preparation, data sharing requests, and initiated the data archives and lesion challenges.

