## Supplementary File for "An updated open-source, large dataset of T1-weighted stroke brain MRIs and lesion segmentation masks"

### Supplementary Material

#### Methods

##### *Data Overview*

**Supplementary Table 1. Cohort-specific metadata on subjects included in the training dataset (n=1453)**

Cohort-specific metadata on the number of subjects included in the public training dataset, median days post stroke, institution and country of data acquisition are provided. Abbreviations: SOOP = Stroke Outcome Optimization Project.

| ***Research Cohort*** | ***Subjects in Training Dataset (n)*** | ***Institution*** | ***Country*** |
|---|---|---|---|
| 1 | 39 | University of Southern California | United States |
| 2 | 12 | University of Southern California | United States |
| 3 | 15 | University of Southern California | United States |
| 4 | 37 | University of Tübingen | Germany |
| 5 | 37 | Emory University | United States |
| 8 | 7 | University of Southern California | United States |
| 9 | 111 | University of Oslo, Oslo University Hospital | Norway |
| 10 | 26 | Tianjin Medical University General Hospital | China |
| 11 | 29 | Tianjin Medical University General Hospital | China |
| 14 | 9 | Medical University of South Carolina | United States |
| 15 | 23 | Medical University of South Carolina | United States |
| 16 | 1 | Medical University of South Carolina | United States |
| 17 | 16 | Medical University of South Carolina | United States |
| 18 | 11 | Medical University of South Carolina | United States |
| 19 | 14 | University of Greifswald | Germany |
| 20 | 1 | University of Greifswald | Germany |

| | | | |
|---|---|---|---|
| 23 | 15 | University of Sciences, Philadelphia | United States |
| 24 | 21 | Emory University School of Medicine | United States |
| 27 | 32 | University of Maryland, Baltimore | United States |
| 28 | 26 | University of California, Irvine | United States |
| 29 | 10 | University of California, Irvine | United States |
| 30 | 15 | University of California, Irvine | United States |
| 31 | 37 | University of California, Irvine | United States |
| 32 | 37 | University of California, Irvine | United States |
| 33 | 2 | University of California, Irvine | United States |
| 34 | 24 | University of Southern California | United States |
| 35 | 15 | Hospital Israelita Albert Einstein, São Paulo | Brazil |
| 38 | 94 | IRCCS Santa Lucia Foundation | Italy |
| 39 | 4 | IRCCS Santa Lucia Foundation | Italy |
| 40 | 86 | IRCCS Santa Lucia Foundation | Italy |
| 41 | 3 | The Florey Institute of Neuroscience and Mental Health<br><br>University of Melbourne | Australia |
| 42 | 35 | Universidad de São Paulo | Brazil |
| 44 | 4 | Medical University of South Carolina | United States |
| 45 | 4 | University of Southern California | United States |
| 46 | 13 | University of Southern California | United States |
| 47 | 48 | University of South Australia | Australia |
| 48 | 44 | University of British Columbia | Canda |
| 49 | 24 | The University of Texas at Austin | United States |
| 50 | 15 | The University of Texas at Austin | United States |
| 52 | 32 | Medical University of South Carolina | United States |
| 53 | 5 | Imperial College London | United Kingdom |

| | | | |
|---|---|---|---|
| 56 | 15 | Casa Colina Hospital and Centers for Healthcare Research Institute | United States |
| 57 | 14 | University of British Columbia | Canda |
| 58 | 35 | Emory University School of Medicine | United States |
| 59 | 38 | Medical University of South Carolina | United States |
| 61 | 15 | University of Southern California | United States |
| 62 | 8 | New York University | United States |
| 63 | 1 | Universitat de Barcelona (UB) | Spain |
| 64 | 6 | Medical University of South Carolina | United States |
| 65 | 18 | Cornell University School of Medicine | United States |
| 67 | 27 | University of Greifswald | Germany |
| 69 | 40 | Casa Colina Hospital and Centers for Healthcare Research Institute | United States |
| 70 | 8 | Emory University School of Medicine | United States |
| 71 | 26 | New York University | United States |
| SOOP | 169 | Prisma Health-Upstate, Greenville | United States |

**Supplementary Figure 1. Cohort-specific metadata on image acquisition parameters (where available) for subjects included in the training dataset (n=1453)**

Cohort-specific metadata on image acquisition parameters for subjects in the training dataset; complete data across parameters was available from 10 research cohorts. Abbreviations: PE = Phase Encoding, MPRAGE: magnetization-prepared rapid gradient-echo.

| Research Cohort | Subjects (n) | Base Resolution | Flip Angle (°) | Echo Time | Field Strength (T) | Manufacturer | Scanner Model | Repetition Time | Acquisition Matrix (PE) | Sequence |
|---|---|---|---|---|---|---|---|---|---|---|
| 5 | 37 | 256 | 8 | 0.00289 | 3 | Siemens | PrismaFit Tim Trio | 2.3 | 256 | MPRAGE |
| 14 | 9 | 256 | 9 | 0.00226 | 3 | Siemens | Tim Trio | 1.9 | 256 | MPRAGE |
| 15 | 23 | 256 | 9 | 0.00226 | 3 | Siemens | Tim Trio | 1.9 | 256 | MPRAGE |
| 16 | 1 | 256 | 9 | 0.00226 | 3 | Siemens | Tim Trio | 1.9 | 256 | MPRAGE |
| 17 | 16 | 256 | 9 | 0.00226 | 3 | Siemens | Tim Trio | 1.9 | 256 | MPRAGE |
| 18 | 11 | 256 | 9 | 0.00226 | 3 | Siemens | Tim Trio | 1.9 | 256 | MPRAGE |
| 24 | 21 | 256 | 9 | 0.00418 | 3 | Siemens | Tim Trio | 2.25 | 256 | MPRAGE |
| 44 | 3 | 256 | 8 | 0.00226 | 3 | Siemens | PrismaFit | 2.3 | 256 | MPRAGE |
| 49 | 24 | 160 | 8 | 0.00416; 0.00417; 0.00419 | 3 | Siemens | Skyra | 2.5 | 160 | MPRAGE |
| 50 | 14 | 192 | 8 | 0.00267 | 3 | Siemens | Skyra | 1.7; 1.8; 1.82; 2; 2.5 | 180; 192 | MPRAGE |

**Supplementary Table 2. Cohort-specific metadata on T1w image voxel dimensions of subjects included in the training dataset (n=1453)**

Cohort-specific metadata on the number of subjects with all 3 T1w image voxel dimensions above 1mm, at least one T1w voxel dimension above 1mm and all 3 T1w image voxel dimensions 1mm (isotropic) are provided. The Stroke Outcome Optimization Project (SOOP) data had 128 subjects with all 3 dimensions as ultra-high resolution (i.e. <1mm$^3$); 37 subjects had at least 1 ultra-high-resolution dimension (i.e. <1mm).

| Research Cohort | No. of subjects with all 3 dimensions >1 mm | No. of subjects with at least 1 dimension >1 mm | No. of subjects with all 3 dimensions =1 mm (isotropic) |
|---|---|---|---|
| 1 | 0 | 0 | 39 |
| 2 | 0 | 0 | 12 |
| 3 | 0 | 0 | 15 |
| 4 | 0 | 2 | 35 |
| 5 | 0 | 0 | 37 |
| 8 | 0 | 3 | 0 |
| 9 | 0 | 5 | 1 |
| 10 | 0 | 0 | 26 |
| 11 | 0 | 3 | 23 |

| | | | |
|---|---|---|---|
| 14 | 0 | 0 | 9 |
| 15 | 0 | 0 | 0 |
| 16 | 0 | 0 | 1 |
| 17 | 0 | 0 | 16 |
| 18 | 0 | 0 | 11 |
| 19 | 0 | 0 | 0 |
| 20 | 0 | 0 | 0 |
| 23 | 0 | 0 | 0 |
| 24 | 0 | 7 | 3 |
| 27 | 0 | 13 | 0 |
| 28 | 0 | 0 | 26 |
| 29 | 0 | 0 | 10 |
| 30 | 0 | 0 | 15 |
| 31 | 0 | 0 | 37 |
| 32 | 0 | 0 | 37 |
| 33 | 0 | 0 | 2 |
| 34 | 0 | 0 | 24 |
| 35 | 0 | 0 | 0 |
| 38 | 0 | 0 | 92 |
| 39 | 0 | 4 | 0 |
| 40 | 0 | 8 | 62 |
| 41 | 0 | 1 | 2 |
| 42 | 0 | 13 | 14 |
| 44 | 0 | 1 | 3 |
| 45 | 0 | 0 | 0 |
| 46 | 0 | 0 | 0 |
| 47 | 0 | 48 | 0 |
| 48 | 0 | 0 | 44 |
| 49 | 24 | 24 | 0 |
| 50 | 14 | 15 | 0 |
| 52 | 0 | 0 | 32 |
| 53 | 0 | 0 | 5 |
| 56 | 0 | 15 | 0 |
| 57 | 0 | 0 | 14 |
| 58 | 0 | 0 | 0 |
| 59 | 0 | 2 | 35 |
| 61 | 0 | 0 | 15 |
| 62 | 0 | 0 | 8 |
| 63 | 0 | 0 | 0 |
| 64 | 0 | 0 | 4 |
| 65 | 0 | 0 | 9 |
| 67 | 0 | 4 | 0 |
| 69 | 0 | 40 | 0 |
| 70 | 0 | 0 | 8 |
| 71 | 0 | 1 | 25 |

| SOOP | 0 | 36 | 0 |
|---|---|---|---|

**Supplementary Table 3. Cohort-specific lesion metadata on subjects included in the training dataset (n=1453)**

Cohort-specific lesion metadata including the median days post stroke, and lesion volumes (in MNI space) for the subjects included in the public training dataset, are provided. As noted in the *Data Overview* section, Chronicity was used when information on days post stroke was not made available by the research cohort. Abbreviations: SOOP = Stroke Outcome Optimization Project.

| *Research Cohort* | *Median Days Post Stroke (Days)* | *Median Lesion Volume (MNI space) (mm$^3$)* |
|---|---|---|
| 1 | 704 | 6160 |
| 2 | 2196 | 31,512 |
| 3 | 1464 | 22,203 |
| 4 | 378 | 51,110 |
| 5 | 1012 | 2111 |
| 8 | 1 | 1631 |
| 9 | 426 | 1419 |
| 10 | 1 | 851 |
| 11 | 1 | 1178 |
| 14 | Not Available | 178 |
| 15 | 1 | 27,148 |
| 16 | Not Available | 1044 |
| 17 | 730 | 20,267 |
| 18 | 836 | 53,090 |
| 19 | 31.5 | 2938.50 |
| 20 | 8 | 5937 |
| 23 | 1 | 67,662 |
| 24 | 518.5 | 5,632 |
| 27 | 610 | 15,643.50 |
| 28 | 575.5 | 7,031.50 |
| 29 | 326 | 5,302.50 |

| | | |
|---|---|---|
| 30 | 545 | 24,464 |
| 31 | 138 | 10,196 |
| 32 | 128 | 7,494 |
| 33 | 156 | 24,908 |
| 34 | 1,506.5 | 3,284 |
| 35 | 1,006.5 | 3,895 |
| 38 | 100 | 6,625.50 |
| 39 | 5.5 | 1,894.50 |
| 40 | 74 | 25,715 |
| 41 | 89 | 235 |
| 42 | 728 | 14,604 |
| 44 | 1,311.5 | 22,103.50 |
| 45 | 2,882.2 | 8,075.50 |
| 46 | 2,589 | 4,888 |
| 47 | 1,024 | 13,357.50 |
| 48 | 1,404 | 11,124 |
| 49 | 0 | 1,278 |
| 50 | 0 | 381 |
| 52 | 1,180.5 | 7,564.50 |
| 53 | 25 | 199 |
| 56 | 555 | 2,813 |
| 57 | 1,703.5 | 29,702.50 |
| 58 | 22 | 5,897 |
| 59 | Not Available | 8,239 |
| 61 | 3,027 | 11,551 |
| 62 | 1,356 | 18,409 |
| 63 | 208 | 5,557 |
| 64 | 1566 | 14,903.50 |
| 65 | 2 | 2,079 |
| 67 | 488 | 21,708 |
| 69 | 20 | 12,551 |
| 70 | 21 | 5688 |
| 71 | 25 | 4182 |

| | | |
|---|---|---|
| SOOP | 180 | 31,309 |
